# Uncontrolled glycemia and PTSD in diabetic patients living in high conflict zones: A cross-sectional study

**DOI:** 10.1101/2024.06.03.24308384

**Authors:** Mohammad Bleibel, Bilal Nasser, Lana El Dorra, Leya Al Jomaa, Hiba Deek

## Abstract

**Background:** Local conflicts such as those occurring in Palestinian camps in Lebanon have led to detrimental effects on the physical and psychological aspects of the people living in these regions. PTSD and uncontrolled glycemia are expected to be the consequences of these events.

**Aim:** To determine the impact of the Ein El Helwe events on blood glycemic levels and the possibility of developing post-traumatic stress disorders.

**Design:** A cross-sectional questionnaire with self-administered questionnaires.

**Methods:** Participants were identified from the community of South Lebanon in the area surrounding, or inside, the Ein El Helwe Camp during the time of the events. The participants completed an electronic questionnaire which included sociodemographic data, diabetes condition and the DSM-IV symptoms of PTSD and its level. The Scale is a validated scale that had previously been translated to Arabic.

**Results:** A total of 234 participants were included in the final analysis of the study with almost 30% having uncontrolled glycemia. The mean age of the study participants was 53.49+13.09 years with more female than male participants and more living around rather than inside the camp. Glycemic control was associated with gender, educational level, living in or outside the camp and the body mass index. Multivariate analysis confirmed the impact of educational level, living area and PTSD on the glycemic control. The relationship between glycemic control and PTSD was based on some of the latter’s symptoms.

**Conclusion:** The study highlighted the impact of conflicts and wars on the development of uncontrolled glycemia among participants living in and around Ein El Helwe Camp. Multiple factors contributed to the findings including sociodemographic, lifestyle factors and cultural aspects which should further be analysed in future studies. Additionally, glycemia levels should be monitored for more reliable findings in future studies.

## Introduction

Lebanon has witnessed a number of instabilities in the past years. These instabilities have impacted the social, physical and psychological wellbeing of individuals residing in Lebanon (1). Lebanon has now been ranked as a low-middle income country following the named crises on many levels (2). Therefore, low socioeconomic status (LSS) is common and increasing in many regions of the country. Ein El Helwe is a camp located in the South Region of Lebanon encompassing many different groups of the Palestinian nationality. These groups are either agreeable together or in conflict with one another (3). This conflict escalated in August 2023 between two groups that lead to a large number of immigrations from the camp and an increase in the death and injured toll. When LSS is combined with post-traumatic stress disorder (PTSD), they were reported to increase the risk of hyperglycaemia in diabetes mellitus type 2 (DMT2) patients (4). In addition, living in high-conflict areas exacerbates the symptoms of PTSD and DM (5).

Post-traumatic stress disorder (PTSD) is group of physical and psychological symptoms that a person manifests secondary to an out of the ordinary event (6). For the traumatic effect to occur a person has to be subjected to an actual, potential, or imagined threat that leaves him/her feeling unable to react, helpless, fearful, and left alone (7). PTSD causes the victim to be in a constant state of hyper-arousal due to the dysregulation of the hypothalamic-pituitary-adrenal (HPA) axis, in return this leads to elevated levels of cortisol; a hormone responsible for maintaining metabolism, blood pressure, and blood sugar levels regulation. In addition to the possible nightmares or repeated visions of the events that has caused the state of stress, anxiety and fatigue are also common (8). According to WHO the PTSD prevalence worldwide was reported by a weighted 70.4% (9). The lifetime prevalence of PTSD in the population was 3.9% with significant variation across countries (10). Although under-reported, psychological conditions are also evident in the Lebanese society and specifically among the refugees residing in camps. These conditions are a result of ongoing battles and conflicts in the camps specifically and in the country in general. In fact, depressive symptoms were noted in more than 50% of the participants of a study conducted between 2017 and 2019 (11). PTSD was also a notable psychologic condition prevalent in the Lebanese society. A recent literature review reported rates ranging between 2% and 98% of PTSD in Lebanon with the variations in these percenatges due to the areas of the studies conducted (under military instability), dates (period since last nearby conflict) and association to those influenced directly by instability (12). Other variables contributing to PTSD were female gender, financial challenges, lower education and number of previous traumas (12).

Gender was seen as a risk factor for most psychological conditions in most of the reviewed studies. In fact, female gender was associated with twice the risk of acquiring PTSD in comparison to male gender (13). This risk factor was also seen among Syrian refugees in north Lebanon where out of 450 participants of the study more than 80% were found to have PTSD symptoms and female gender was of significance (14). Another study looked at 625 citizens residing in south Lebanon also showed PTSD rates of 33% and depression rates of 19%. Female gender was also of significance along with being married, having more children, unemployment, lower educational levels and having witnessed war (15).

Diabetes is described as a chronic state of increased blood glucose levels above normal values (16). It is on the rise worldwide, with a global prevalence in adults in 2017 being 8.8% of the world population, with the anticipation of a further increase to 9.9% by 2045 (17). As of 2017, the prevalence of diabetes in Lebanon was estimated to be 14.6% (18). Stress-induced hyperglycemia (SIH) happens in patients that experience a major traumatic event in addition to multiple physical or life style risk factors including intensive care units admissions (19). SIH has been found to double the rate of mortality risk (20) and that is more true in hospitalized patients (19). The development of PTSD among patients with diabetes can be further emphasised by insulin resistance, uncontrolled glycemia, dysfunction in the hypothalamic-pituitary-adrenal (HPA) axis and psychological contributions (21). PTSD and hyperglycemia can affect the patient’s mental and physical health, as they are related conditions that have a bidirectional relationship. It was reported that stress could peak blood glucose levels, similarly to the levels after completing a meal. Stress, on the other hand, is caused by flawed insulin sensitivity and glucose reuptake, which are associated with diabetes mellitus type 2 (DMT2) (22). The link between these two conditions was established in previous studies among Asylum seekers in the Netherlands. The findings revealed that people with PTSD were more likely to have DM than those without (23). A similar link was seen between PTSD, DM and metabolic syndrome (24). Self-care reflected by glycemic control was evident among those free of psychological problems. This was shown when patients with PTSD or depression were 48% more likely to be in poor glycemic control (25).

Having mentioned the effect of traumas and conflicts on the physio-psychosocial wellbeing of individuals, it was necessary to study the effect of the named conflicts in South of Lebanon and specifically in the Ein El Helwe camp on physical and psychological wellbeing of people residing in Saida region. Therefore, the aim of the current study was to evaluate the effect of Ein El Helwe events on the development of PTSD and hyperglycemia among diabetic patients living in Saida community at the time of Ein El Helwe events.

## Methods

### Design

A cross-sectional design involving self-administered questionnaires and interviews.

### Settings

Saida community.

### Eligibility Criteria

The research was conducted on community residents in Saida following Ein el Helwe events in August 2023. Participants aged more than 18 years and were diagnosed with diabetes mellitus types 1 and 2 were invited to participate. Those with other types of diabetes such as gestational diabetes were excluded. Gestational diabetes and steroid diabetes were excluded for the possibility of their remission after the cause of the presence is interrupted and thus lacking the chronicity of the condition (26).

### Procedure

Ethical approval was secured from the Institutional Review Board at Beirut Arab University under the approval code of 2023-H-0157-HS-M-0551. A link with the Arabic version of the data collection form was generated on Google Forms and was disseminated among Saida residents through Social Media platforms and through face-to-face. Consent were taken from those interviewed verbally whereas those submitted answers on google forms were asked whether they agree or not to complete the survey. Data collection took place over a two-months period with weekly reminders sent out on social media. The data collection form included the following sections:

A. **Sociodemographic Data:** This section included data on age, gender, weight, height, educational level, occupation, nationality, marital status, place of living and monthly income.
B. **Past medical and surgical history:** This section included questions on the diagnosed medical conditions and the surgical history. In addition to that, medication use, and adherence were also tackled to rule out uncontrolled diabetes due to poor medication adherence. To account for the significant and common comorbidities, and to give a weight for each ailment, the Charlson’s comorbidity index was used (27). A calculated weight of the CCI was used in the analysis.
C. **Diabetes (readings-medications-monitoring)**: This section addressed the different variables that may impact blood sugar levels. Questions included diabetes type, current medications with the doses, diet restrictions, personal blood sugar level monitoring at home and the number of times per day, last fasting blood sugar level, last HBA1C level and the last check up with his physician. Additionally, significant symptoms of high glucose levels were also screened. Glycemia targets were set for 180mg/dL where above which was categorized as hyperglycemia and below which was controlled glycemia among diabetic patients (28).
D. **Post-Traumatic Stress Disorder scale (PTSD):** This 17-item scale evaluates the DSM-IV symptoms of PTSD and its level. The items are rated on a five-point Likert scale ranging from 1 (never) to 5 (always) with total numbers ranging between 17 and 85 and higher scores indicate severer forms of PTSD (29). The items of the PTSD scale reflect the symptoms of this condition where items 1 to 5 reflect re-experiencing (B Symptoms) with a score range between 5 and 25. Items 6 to 12 address avoidance/numbing symptoms (C symptoms) with scores ranging between 5 and 35. The third construct addresses hyperarousal (D symptoms) through items 13 to 17 with scores ranging between 5 and 25. Diagnosis based on DSM-IV symptoms of PTSD was determined from the symptom clusters where at least one B symptoms, three C symptoms and two D symptoms were considered a positive PTSD diagnosis (30). The Arabic version of the tool has been used previously and evaluated for psychometric properties and shown to have a Cronbach alpha of 0.89 for the total score. Additionally confirmatory factor analysis results showed adequate fit of the critical values to the model (29).

### Sample size calculation

This study was found to be replicating another with a similar outcome (31). The primary outcome is PTSD among patients with diabetes or hyperglycemia in the Lebanese South Region. Therefore, a similar sample size was sought for the current study which 220 participants.

### Data analysis

Data were imported from excel sheet and analyzed using the Statistical Product and Service Solutions (SPSS) version 24. Continuous variables such as (age-height) were presented as means and standard deviations whereas, categorical variables were presented as frequencies and percentages. Group comparison was based on glycemic control with a target of 180mg/dL (28). The other group comparison was based PTSD scoring according to the DMS-IV symptoms of PTSD (30). The Chi square test and t-test were used for bivariate analysis based on the level of data. Multiple logistic regression was done to identify a model with the predictors of controlled glycemia at the multivariate level. The variables were derived from the literature and those that were significant at the bivariate level. Significance was set for a p value of less than 0.05.

## Results

### Sociodemographic characteristics and medical profile of the study participants

A total of 234 participants were included in the final analysis of the study. The mean age was 53.49 (SD=13.99) years, and the majority were female participants (n=125, 53.4%). The mean weight and height were 79.52 (SD=11.43) kg and 170.2 (SD=7.04) cm respectively with a calculated body mass index (BMI) of 27.44 (SD=3.88). The majority were married (n=171, 73.1%), and the educational level was mostly university level accounting for 35% of the sample. Two-fifths were unemployed (n=99, 42.3%), the majority lived around Ein El Helwe camp (n=182, 77.8%) and two-thirds were Palestinians (n=151, 64.5%). Almost two-thirds of the sample were smokers (n=144, 61.5%) and only 5.6% (n=13) consumed alcohol. The mean income/month was $378.19 (SD=460.51) U.S. In terms of medical conditions, almost two-thirds had chronic conditions besides diabetes (n=140, 59.8%) with half having HTN (n=121, 51.7%), less having hypercholesterolemia (n=90, 38.5%), and only 5.1% having COPD (n=12). Details about the sociodemographic characteristics and the medical profile of the study participants are presented in Table 1.

**Table 1:** Sociodemographic characteristics and medical profile of the study participants (N=234)

| Variables | Total<br>(N=234, 100%) |
| --- | --- |
| Age* | 53.49 (13.99) |
| Gender |  |
| • Female | 125 (53.4) |
| • Male | 109 (46.6) |
| Weight* | 79.52 (11.43) |
| Height* | 170.2 (7.04) |
| BMI | 27.44 (3.88) |
| Social status |  |
| • Married | 171 (73.1) |
| • Divorced/widowed | 41 (17.5) |
| • Single | 22 (9.4) |
| Educational Status |  |
| • University level | 88 (35) |
| • High school level | 79 (33.8) |
| • Elementary school | 49 (20.9) |
| • Not educated | 24 (10.3) |
| **Occupation** |  |
| • Unemployed | 99 (42.3) |
| • Full time | 84 (35.9) |
| • Part Time | 51 (21.8) |
| **Living Area** |  |
| • Around Camp | 182 (77.8) |
| • In Camp | 52 (22.2) |
| **Nationality** |  |
| • Palestinian | 151 (64.5) |
| • Lebanese | 79 (33.8) |
| • Syrian | 4 (1.7) |
| **Smoker** | 144 (61.5) |
| **Alcohol** | 13 (5.6) |
| **Income/ month \$\*** | 378.19 (460.51) |
| **Chronic medical conditions** | 140 (59.8) |
| **HTN** | 121 (51.7) |
| **Hypercholesterolemia** | 90 (38.5) |
| **Skin ulcer/ cellulitis** | 1 (0.4) |
| **Any tumor** | 2 (0.9) |
| **Moderate & severe liver disease** | 1 (0.4) |
| **Mild liver disease** | 2 (0.9) |
| **Hemiplegia** | 1 (0.4) |
| **Ulcer disease** | 5 (2.1) |
| **MI** | 48 (20.5) |
| **Renal disease** | 6 (2.6) |
| **PVD** | 21 (9) |
| **COPD or asthma** | 12 (5.1) |
| **Rheumatic disease** | 18 (7.7) |
| **Depression** | 5 (2.1) |
| **CHF** | 26 (11.1) |
| **DM with organ damage** | 7 (3) |
| **On warfarin** | 9 (3.8) |
| **Lymphoma** | 1 (0.4%) |

Legend: HTN: hypertension; PVD: peripheral vascular disease; COPD: chronic obstructive pulmonary disease; CHF: congestive heart failure; MI: myocardial infarction; DM: diabetes mellitus
\*:data presented in frequencies, means and standard deviation;

### Characteristics of diabetes condition and lifestyles among the study participants

The majority of the participants had type 2 diabetes (n=216, 92.3%). The mean years since diagnosis was 7.09 (SD=7.56) and more than three-fourths were on metformin (n=180, 76.9%). Most of the participants were following a specific diet for diabetes (n=173, 73.9%) and regularly evaluated their hemoglobin-A1C (HBA1C) (n=228, 97.4%). Almost two-thirds had their HBA1C within normal ranges of 4-5.5 (n=144, 61.5%) while 29% had an elevated HBA1C of more than 5.5 (n=68). In terms of frequent home monitoring, one-third took their hemo-glucose test (HGT) once daily (n=77, 32.9%) with a mean fasting hemo-glucose test of 141.26 (SD=51.39) mg/dl. In terms of control, 33 (15%) had fasting glycemia levels above 180mg/dL as reported by the patients. Despite that, a large number complained of excessive thirst (n=184, 78.6%), polyuria (n=201, 85.9%), exhaustion (n=172, 73.5%), blurred vision (n=149, 63.7%), numbness (n=173, 73.9%), and excessive hunger (n=135, 57.7%). Finally, two-thirds had regular check-ups with their physicians (n=158, 67.5%). Details about the diabetes conditions and lifestyles are presented in Table 2.

**Table 2:** Characteristics of diabetes condition among the study participants (N=234).

| Variables | Total (234, 100%) |
| --- | --- |
| **DM types** |  |
| • Type 2 | 216 (92.3) |
| • Type 1 | 18 (7.7) |
| **Diagnosis since\*** | 7.09 (7.56) |
| **Amaryl** | 1 (0.4) |
| **Avandia** | 1 (0.4) |
| **Diamicron** | 6 (2.6) |
| **Galvus met** | 22 (9.4) |
| **Insulin** | 18 (7.7) |
| **Jardiance** | 1 (0.4) |
| **Kazano** | 1 (0.4) |
| **Metformin** | 180 (76.9) |
| **Sulfonylurea** | 1 (0.4) |
| **Trajenta** | 2 (0.9) |
| **Specific diet for DM** | 173 (73.9) |
| **Do you take your HBA1C** | 228 (97.4) |
| **What was the last average?** |  |
| • 4-5.5 | 144 (61.5) |
| • >5.5 | 68 (29.1) |
| • <4 | 22 (9.4) |
| **How many times do you test your hemo-gluco test?** | 77 (32.9) |
| • Once daily | 60 (25.6) |
| • Once every two days | 48 (20.5) |
| • Rarely or never | 35 (15) |
| • Once a week | 14 (6) |
| • Twice daily |  |
| **Average hemo-gluco test\*** | 141.26 (51.39) |
| **Excessive thirst?** | 184 (78.6) |
| **Polyuria** | 201 (85.9) |
| **Exhaustion** | 172 (73.5) |
| **Blurred vision** | 149 (63.7) |
| **Numbness** | 173 (73.9%) |
| **Excessive hunger** | 135 (57.7) |
| **Regular follow ups with physician** |  |
| • Yes | 158 (67.5) |
| • Rarely | 66 (28.2) |
| • No | 10 (4.3) |

Legend: \*:data presented in frequencies, means and standard deviation; \*\*:significant at less than 0.05. DM: Diabetes Mellitus, HBA1C: Hemoglobin A1C (Glycated Hemoglobin).

### Characteristics of the PTSD scale among the study participants

The mean PTSD score was 45.84, SD=9.94 with 158 (71.8%) found to have PTSD based on the DSM-IV symptoms category. The most prevalent symptoms reflecting diagnosis was the reexperiencing symptom occurring in 205 (93.2%) of the study participants.

In terms of group difference, it was noted that the mean score did not differ between those with controlled and uncontrolled glycemia. However, when looking at the PTSD symptom categories of the DMS-IV, it was noted that those with controlled glycemia were significantly more likely to have PTSD in comparison to those with uncontrolled glycemia (143, 90.5% vs. 15, 9.5%); p=0.000). This was reflected in the Symptom C category; avoidance/numbing, as indicated in Table 3.

**Table 3:** PTSD scale scoring of the study participants (n=234).

| Variables | Total<br>(N=234, 100%) | Uncontrolled glycemia<br>(n=33, 15%) | Controlled glycemia<br>(n=187, 85%) | P value |
| --- | --- | --- | --- | --- |
| Total PTSD mean Score* | 45.84 (9.94) | 46.06 (9.88) | 45.78 (10.11) | 0.88 |
| PTSD symptoms categories | 158 (71.8) | 15 (9.5) | 143 (90.5) | 0.000 |
| Re-experiencing symptoms (B) | 205 (93.2) | 30 (14.6) | 175 (85.4) | 0.57 |
| Avoidance/numbing (C) | 165 (75) | 16 (9.7) | 149 (90.3) | 0.000 |
| Hyperarousal (D) | 204 (92.7) | 173 (84.8) | 31 (15.2) | 0.77 |

LEGEND: \*:data presented in frequencies, means and standard deviation; \*\*:significant at less than 0.05.

### Predictors of controlled glycemia among the study participants

Gender, educational status and residence location contributed to glycemia control. This was evident when those with controlled glycemia were significantly more likely to be educated (149, 94.9% vs. 8, 5.1%; p=0.000) and female gender (106, 89.8% vs. 12, 10.2%; p=0.03) than those who were not or ere of male gender. Additionally, this same group was significantly more likely to be resident of the area surrounding the camp in comparison to those living inside he camps (156, 83.4% vs. 31, 16.6%; p=0.00). In terms of lifestyle, it was noted that those with controlled glycemia, were significantly more likely to follow diet regimens that suited their health condition than those without glycemia control (151, 91% vs 15, 9%; p=0.00) respectively. Moreover, those with lower BMI (26.88+3.14 vs. 29.55+5.8; p=0.014) and less years since diagnosis (6.29+6.89 vs. 11.67+10.34; p=0.007) were more likely to have controlled glycemia than their counterparts.

Logistic regression analysis was done to identify the best fitting model for glycemic control. The model included gender, living location, PTSD development and body mass index with the first three remaining significant throughout the analysis. The overall model was significant (*x*^2^=55.295, df=4, p=0.00) with a good fit and a non-significant Hosmer and Lemshow test (x2=11.04, df=8, p=0.199). The model was able to classify 89.3% of the sample with more in the controlled glycemia group (96.7%) than in the uncontrolled glycemia group (48.5%). The variance explained by the model was moderate to account to almost 40%. Details about the logistic regression analysis are presented in Table 4.

**Table 4.** Predictors of controlled glycemia using logistic regression (N=215)

| Variables | B | S.E | Wald | df | Sig | Exp (B) | 95% C.I |  |
| --- | --- | --- | --- | --- | --- | --- | --- | --- |
|  |  |  |  |  |  |  | Lower | Upper |
| **Gender** | 0.889 | 0.473 | 3.531 | 1 | 0.06 | 2.432 | 0.962 | 6.148 |
| **Education level** | 2.246 | 0.488 | 21.208 | 1 | 0.000 | 9.454 | 3.634 | 24.594 |
| **Living area** | 1.313 | 0.470 | 0.470 | 1 | 0.005 | 3.717 | 1.481 | 9.331 |
| **BMI** | 0.095 | 0.056 | 2.921 | 1 | 0.087 | 1.100 | 0.986 | 1.227 |
| **PTSD** | 1.114 | 0.471 | 5.591 | 1 | 0.018 | 3.046 | 1.211 | 7.664 |
| **constant** | -6.345 | 1.671 | 14.424 | 1 | 0.000 | 0.002 |  |  |

Legend: S.E: standard error of the coefficient; df: degree of freedom; Exp: exponential; CI: confidence interval; BMI: Body mass index; PTSD: post-traumatic stress disorder.

## Discussion

The main aim of the study was to evaluate the impact of Ein El Helwe events on blood glycemia level and the possibility of developing post-traumatic stress disorders among patients with known diabetes among residents of the South Region of Lebanon. This was evaluated using a cross-sectional approach using a self-administered questionnaire. The main results revealed that one third of diabetic patients had no glycaemic control reflected by a reported fasting glycemia level of above 180mg/dL. This percentage was lower than that found in the Literature where hyperglycaemia was noted in more than 50% of the patients with diabetes. The latter study was aimed to evaluate the health-related quality of life of the sample living in Stockholm (32). The higher levels could be attributed to the higher mean age of the sample which was 73.80 years in comparison to the younger ages in our sample of a mean age of 53.49 years. Slightly more recent studies showed almost similar percentages of 45% of patients with uncontrolled glycemia among diabetics (33). However, much higher rates of uncontrolled glycemia to almost 80% were noted in a most recent study. Despite the comparable age groups with the current study, the high rates could be reflecting the challenges experienced by the study population of Iran with the political and social changes they are manifesting. Additionally, the time of the study coincided with the COVID-19 era, which imposed additional challenges on the populations (34).

The other aim was to evaluate PTSD level among diabetic patients in Saida region which was evaluated to be 45.85 (SD=9.94) reflecting moderate PTSD. When looking further into the results it was noted that only 6 (2.6%) had no symptoms associated with PTSD while almost two-thirds had more than three symptoms of PTSD reflecting severity. PTSD was reported to be prevalent in 30% of patients with diabetes due to many variables including fear of death, feeling of helplessness and HBA1C numbers (35). The latter study showed lower rates of PTSD prevalence due to younger ages of participants having Type 1 diabetes in contrast to the current study of older ages with Type 2 diabetes.

The findings revealed no significant difference in the mean scores of PTSD among those with controlled versus uncontrolled glycemia in the current study. This contradicts previous studies outlining higher rates of PTSD among the diabetics when compared to those without the condition (36). However, no studies were found to compare PTSD among diabetics with and without glycemic control. An interesting notion was the similarity in the scoring of item number 10 of the scale between those with and those without controlled glycemia. This item addressed feeling cut off from society which can be attributed to the type of culture Arabic people in general adopt. This is the collectivism which proves to be an encompassing environment of all its individuals (37). The fact that PTSD has an impact on people of outstanding traits within their culture i.e. individualists in collectivist societies (38), needs to be further investigated in future studies to outline the actual source and rates of this psychologic condition in the presence of chronic conditions.

Although the sample size fells within ranges of those found in the literature with similar objectives (39), the main limitation of the study was the poor generalizability of the sample to non-married individuals as less than 30% were single or divorced/widowed. This has an impact on the PTSD development as the Arab culture is known be a collectivist culture where social support is naturally provided, thus decreasing the incident of PTSD development (40). However, the sample was indeed generalizable for the South region where the conflicts had been happening and the genders which were equally distributed. Another generalizability concern is the type of diabetes which was much higher among the type 2. This should be addressed in future studies by restricting the types to either type 1 or 2 for a better insight. Another limitation is the approach of the study which was cross-sectional and depending on the reporting of the participants of their diabetes conditions, glycemic control and follow up. This could have produced response bias due to social desirability effect. Future studies should include glycemia monitoring of the study participants for more reliable results.

## Conclusion

The study aimed to assess the impact of Ein El Helwe incidents on blood glucose levels and the likelihood of developing post-traumatic stress disorder following the events. The results showed that a significant number had controlled glycemia level which came as a surprise considering the social and political challenges this cohort faced. However, an important element to consider is the resilience which the people of Lebanon and especially the Palestinians own. This should be considered in future studies to evaluate the effect of these variables on glycemia level. Another future consideration is the design of the study which should include direct evaluation on the glycemic level rather than depending on the reported numbers and conditions of the study participants. Despite all, the study brings important insights on the diabetes and lifestyle practices that people in highly challenged living conditions follow to maintain their wellbeing. The study also highlights the need for education to be provided to patients with diabetes about these lifestyle measures to enhance better control of their glycemic levels.

## Data Availability

All relevant data are within the manuscript

## Acknowledgment

The authors of this paper would like to express their gratitude to the Saida community for their assistance in completing the questionnaire for this study.

